# US Life Expectancy Rebounds in 2022

**DOI:** 10.1101/2023.02.26.23286363

**Authors:** Harry Wetzler

## Abstract

From 2019 to 2021 life expectancy at birth in the United States (US) declined by approximately 2.45 years to values not seen since 1996. Complete life tables for 2019 to 2022 were constructed for the total US population, females, and males using mortality data from the CDC WONDER Multiple Cause of Death database and Census Bureau Vintage population estimates. Life expectancy at birth increased by 1.07 years between 2021 and 2022. Nearly 40% of lost life expectancy years at birth were regained in 2022.

## Introduction

US life expectancies at birth declined by 2.1 years for females and 2.8 years for males between 2019 and 2021.^1,2^ The resultant life expectancies in 2021 were the lowest seen since 1996.

Multiple Cause of Death data in the CDC WONDER database are updated monthly.^3^ The February 5, 2023 data release was combined with Vintage Census Bureau population estimates to construct preliminary 2021 and 2022 life tables for the total, female, and male US populations. Life tables for 2019 and 2020 were similarly constructed for comparison with the published life tables for those years.^4,5^

## Methods

Death data for 2019-2022 by year, gender, and single year of age were obtained from the CDC WONDER Multiple Cause of Death database as of Jan 21, 2023.^6^ Since the 2022 death data are provisional and incomplete, monthly total death changes beginning in July 2022 and ending in January 2023 were modelled to develop inflation factors for the previous eleven months. A cubic polynomial fits the average monthly changes very well (r-squared=0.962). Although there is evidence that death certificate reporting timeliness varies by age and sex, a common monthly inflation factor was used because the monthly changes by age and sex were not available for this study.^7^ Population data for July 1, 2019 were downloaded from the Census Bureau Vintage 2020 population estimates and data for July 1, 2020-2022 from the Vintage 2021 estimates.^8,9^ Standard life table techniques were used to construct complete life tables for 2019-2022.^5^ Data for life years lived in the first (age 0 to 1) and last (age 100+) intervals for 2019-2020 were obtained from published life tables.^4,5^ First and last year interval data from 2020 were used in the new 2021 and 2022 life tables constructed for this study. Life expectancy estimates obtained from WONDER death and Vintage population data for 2019-2021 were compared to published values from the Centers for Disease Control (CDC) and the National Center for Health Statistics (NCHS). Chiang’s formula was used to calculate the standard error of life expectancy.^10^

## Results

The CDC Wonder Multiple Cause of Death database tallied 3,251,106 deaths in 2022 as of Jan 21, 2023. A total of 3,310,221 deaths are forecast to occur in 2022 resulting in an overall inflation factor of 1.0182. Table 1 lists life expectancies at birth for the published US life tables and those calculated in this study.

|  |  | Year |  |  |  |
| --- | --- | --- | --- | --- | --- |
|  |  | 2019 | 2020 | 2021 | 2022 |
| Total | CDC/NCHS | 78.85 | 76.99 | 76.4 | - |
|  | This study | 79.13 | 77.03 | 76.41 | 77.48 |
| Female | CDC/NCHS | 81.39 | 79.88 | 79.3 | - |
|  | This study | 81.56 | 79.83 | 79.34 | 80.18 |
| Male | CDC/NCHS | 76.31 | 74.19 | 73.5 | - |
|  | This study | 76.57 | 74.34 | 73.60 | 74.87 |

**Table 2.** lists life expectancies at age 65 for the published US life tables and those calculated in this study.

|  |  | Year |  |  |  |
| --- | --- | --- | --- | --- | --- |
|  |  | 2019 | 2020 | 2021 | 2022 |
| Total | CDC/NCHS | 19.59 | 18.46 | 18.40 | - |
|  | This study | 19.92 | 18.42 | 18.48 | 18.93 |
| Female | CDC/NCHS | 20.82 | 19.78 | 19.70 | - |
|  | This study | 21.01 | 19.69 | 19.77 | 20.12 |
| Male | CDC/NCHS | 18.20 | 17.03 | 17.00 | - |
|  | This study | 18.52 | 17.04 | 17.08 | 17.63 |

Inspection of Table 1 reveals that the differences between published and newly calculated life expectancies decreased from 2019 to 2021, averaging -0.05 years (published value minus new value; note that since the published 2021 life expectancies are only accurate to tenths of a year, the differences could actually range from -0.10 to 0.00) in 2021. In terms of years of life expectancy at birth lost between 2019-2021, approximately 40% of those years were regained in 2022. The increase in years of added life expectancy for 2022 compared to 2021 was 51% more for males than females. Yet it is important to note that years of life expectancy at birth declined by 35% more for males than females between 2019 and 2021. The standard error of 2022 life expectancy at birth was 0.0085 years.

Again, the differences between published and new values decreased after 2019 with an average difference in 2021 of -0.08 years. Fewer years of lost life expectancy at age 65 were regained from 2021 to 2022 and males regained more, 38.2%, than females, 28.2%. Note that years of life expectancy at age 65 declined by only 7% more for males than females between 2019 and 2021.The standard error of 2022 life expectancy at age 65 was 0.0053 years.

## Discussion

After declining for two years, US life expectancy reached a nadir in 2021 before rebounding in 2022. However, less than half of the 2019-2021 losses have been made up. With continuing daily COVID-19 deaths exceeding 400 nationwide as of Feb 15, 2023, it is doubtful that the life expectancy decreases from 2019 will be eliminated in 2023.^11^

These analyses are subject to a number of caveats. Foremost is the provisional nature of the 2022 death data. The January 21, 2023 totals were adjusted based on earlier 2022 reporting trends but it is impossible to know the accuracy of the results at this time. If the actual number of deaths in 2022 was 1% more than projected (an additional 33,100 deaths), then 2022 life expectancy at birth would decrease by 0.12 years. In addition, the population denominators are estimates. The Vintage 2021 total for the US population on July 1, 2022 was 332,838,183 but the total on that date according to the US Population Clock was 333,287,557, 0.13% more. We do not know the age and gender breakdown of the difference but larger denominators result in lower death rates and greater life expectancy. However, if the increase is applied uniformly across the ages, the added life expectancy at birth is only 0.017 years. In both instances (death numerators and population denominators), NCHS uses more sophisticated techniques involving, for example, blending vital statistics mortality data with Medicare death data to improve the death rate numerators at older ages. It is possible to estimate life expectancy for other population subgroups such as Hispanics and Non-Hispanic American Indian and Alaskan Natives that were more affected by the COVID-19 pandemic and these may be included in future reports.

## Conclusion

After declining 2.45 years between 2019 and 2021, estimated US life expectancy at birth increased by 1.07 years from 2021 to 2022. Comparable estimates for females are a decline of 2.09 years and an increase of 0.84 years; for males the decline was 2.81 years with an increase of 1.27 years. These are preliminary estimates.

## Data Availability

All data produced in the present work are contained in the manuscript. Source data are in the public domain and available online.

https://wonder.cdc.gov/controller/datarequest/D176

https://www2.census.gov/programs-surveys/popest/datasets/2010-2020/national/asrh/

https://www2.census.gov/programs-surveys/popest/datasets/2020-2021/national/asrh/

